# THE COMMERCIAL PROMOTION OF ELECTRONIC CIGARETTES ON SOCIAL MEDIA AND ITS INFLUENCE ON VAPING INITIATION AND POSITIVE PERCEPTIONS OF VAPING IN ANGLOPHONE COUNTRIES: A SCOPING REVIEW PROTOCOL

**DOI:** 10.1101/2022.11.11.22282178

**Authors:** L Chacon, G Mitchell, S. Golder

## Abstract

**Introduction:** Social media plays a protagonist role in the promotion of addictive substances targeting vulnerable and susceptible groups including adolescents and young adults in contemporary times. Despite the broad evidence of tobacco promotion on popular social media, electronic cigarette advertisement remains a recent public health concern, as is considered an emerging phenomenon. Further research must be developed to address this topic, contributing to improved regulation of the marketing of vaping products.

**Aims and Objectives:** The proposed scoping review aims to clarify key concepts related to e-cigarette promotion on social media, synthesise the relevant evidence related to the topic, and identify gaps that will contribute to further research and policymaking enhancement.

**Identifying Studies:** The databases Embase, MEDLINE, PsycINFO, Epistemonikos, Science Citation Index, Cochrane Database of Systematic (CDSR) and CENTRAL will be used to search for relevant studies by using a range of keywords in the search strategy.

**Study Selection:** Studies must comprise individuals over 10 years old living in core anglophone countries. Eligible studies must evaluate the promotion of e-cigarettes on accessible social media platforms, and its influence on vaping uptake and positive perception among the research population. The screening and extraction stage will be held on Covidence through three independent reviews and results will be displayed via a PRISMA flowchart.

**Reporting the results:** Extracted data will be reported through a table and the results presented using a narrative summary. No assessment for risk bias or formal synthesis will be included as this is a scoping review. The results will provide relevant information and recommendations for prospective research.

## INTRODUCTION

Electronic nicotine delivery systems (ENDS) are a group of products created for users’ aerosol inhalation through the heating of an e-liquid, typically containing chemicals that are toxic to human health (World Health Organization, 2022). Electronic cigarettes, or e-cigarettes, are the most popular type of ENDS, despite e-cigars and e-pipes also being commercially available. Differently from conventional cigarettes, electronic cigarettes do not burn tobacco, but instead, heat a liquid substance by using an electronic device, creating an inhaled vapour. This liquid is usually a combination of nicotine, propylene glycol, and diverse flavours (Hiemstra and Bals, 2016).

Electronic cigarettes were first manufactured in the city of Beijing (China) in 2003, and after a year of production, the company Dragonite International marketed the products (Yamin, Bitton and Bates, 2010). Since its commercial availability, diverse electronic cigarette brands multiplicated across the globe (Bertholon et al., 2013), and the products were usually promoted as an alternative to conventional cigarettes and a smoking cessation aid (Hartmann-Boyce et al., 2021). Considering this main appeal, electronic cigarettes are mostly designed to resemble cigarettes in shape and design and can be disposable or reusable depending on their configuration (Brown and Cheng, 2014).

Evidence suggests that electronic cigarettes are in fact, a healthier alternative to cigarettes as they do not expose users to elevated levels of tobacco-related damaging toxins. However, despite the advantage of containing fewer toxicants than cigarettes (Hartmann-Boyce et al., 2021), the use of electronic cigarettes as a smoking cessation aid is questionable according to the World Health Organization (WHO), which grounds its global tobacco control strategies on evidence-based public health interventions and policies (World Health Organization, 2022).

Widely accepted as a safer option than a conventional cigarette, electronic cigarettes are not free of risks (Marques, Piqueras and Sanz, 2021), as diverse harmful substances are included in their composition, in addition to nicotine. Vaping is a dangerous practice among all age ranges, including children, adolescents, adults, and elderly individuals (Centers for Disease Control and Prevention, 2022). The unsafe substances contained in the electronic cigarette aerosol are inhaled by humans and transported to their respiratory system, also affecting non-vapers through passive contamination (Centers for Disease Control and Prevention, 2022).

Following the introduction of electronic cigarettes in society, the human inhalation of micro-particles and volatile chemicals contained in vape devices caused severe lung complications. After a series of cases were identified and reported, the disease was termed EVALI, recognised in medical terms as e-cigarette or vaping product use-associated lung injury (Belok et al., 2020). Tetrahydrocannabinol and vitamin E acetate were identified as the main substances associated with EVALI incidence (Layden et al., 2020).

Despite the impact on the human respiratory system (Belok et al., 2020), vaping has other health risk implications, affecting, for instance, brain development in adolescents, as this organ is not mature until 25 years old. In addition, the toxicological effect of e-cigarettes’ nicotine component on the human brain can affect learning abilities, control attention (Taylor et al., 2014), and neurological synapse formation (Centers for Disease Control and Prevention, 2022). Despite the lack of longitudinal and large-scale studies assessing the relationship between electronic cigarettes and oral cancer development (Raj et al., 2020), several carcinogenic agents are evidenced in e-cigarette brands (Cheng, 2014).

Electronic cigarettes, beyond their negative health impact, are broadly associated as a gateway for cigarette subsequent use, as evidenced in relevant studies around the globe, including the UK (Conner et al., 2017) and the United States (Barrington-Trimis et al., 2016). Furthermore, youths and young adults primarily exposed to vaping practices have more than triple the odds of experimenting with conventional cigarettes (Soneji et al., 2017). Therefore, electronic cigarettes are considered a predictor of conventional smoking (Adermark et al., 2021), and potential access to illicit drug use such as marijuana (Westling et al., 2022).

The vape industry (e-cigarette industry) focuses on appealing strategies to attract potential new consumers by targeting vulnerable populations including children and adolescents (Cataldo et al., 2015). As observed in conventional tobacco products such as cigarettes (Feirman et al., 2015), electronic cigarettes instigate users’ preferences for more flavoured products, by masking the harshness and bitter taste of nicotine (Kim et al., 2016). Thus, flavoured electronic cigarettes contribute to the increase in vaping practices, by altering users’ risk perceptions (Pepper, Ribisl and Brewer, 2016).

The diversity of electronic cigarette flavours in the market and the constant nomenclature variations difficult the development of studies related to the topic (Yingst et al., 2017), but despite such limitations, evidence suggests that youths are more likely to choose sweet and fruity-flavoured electronic cigarettes instead of tobacco or alcohol flavoured ones. This motivation can be partially explained by youths’ reduced harm beliefs (Pepper, Ribisl and Brewer, 2016) and contributes to understanding how the vape industry target vulnerable individuals such as youths through appealing strategies (Hammal and Finegan, 2016).

The desirability of electronic cigarettes among adolescents is strongly related to social acceptability, as vaping indoors and outdoors is more tolerated in comparison with conventional tobacco (Gorukanti et al., 2017). Positive attitudes towards vaping and peer-to-peer influence are also linked with electronic cigarette use among youths (Barrington-Trimis et al., 2015). Thus, social factors including social norms and social interactions play a relevant role in vaping practices (Amin, Dunn and Laranjo, 2020).

As observed previously in the tobacco industry marketing strategies, cigarette promotion in traditional media such as television, radio, and movies played a significant role in smoking behaviours (Evans et al., 1995). Despite denying their direct involvement with the entertainment business, the tobacco industry benefited from the influence of celebrities, popular movies, accessible magazines, and public events to foment their cigarette brands (Mekemson and Glantz, 2002).

The business success of Marlboro, the most popular cigarette brand in the world contributes to the understanding of marketing strategies derived from the tobacco industries, providing opportunities for more efficient control policies (Hafez and Ling, 2005). One of the main marketing tactics of the tobacco industry used to recruit new consumers is promoting their products in association with wealth, pleasure, power, and fun experiences (Mekemson and Glantz, 2002). Therefore, advertising can be explained as the use of media to connect people with desirable personal traits or promote a positive image of a commercial product (Lovato, Watts and Stead, 2011).

Despite the transition from traditional communications channels to new media platforms, the tobacco industry benefits from online marketing by using a broader public reach range, increased accessibility, sharing recourses, and communications possibilities (Freeman, 2012). Social networking sites popularly known as social media are considered a group of internet-based applications, which allow the creation and interchange of the content generated by their users (Kaplan and Haenlein, 2010). Considering the popularity of social media in the world, the vaping industry is adapting and improving marketing strategies, to access a diverse type of public and correspond to their specific demands and interests (Liang et al., 2015).

One of the distinguished advantages of social media advertisements is related to their capacity to direct promotional content to potential consumers based on users’ demographic characteristics and previous online search records (Kaplan and Haenlein, 2010). Additionally, social media generates promotional content by using its audience as an interactive and engagement tool, as users can disseminate the advertising information within their social network, increasing product exposure and consumer range (Camenga et al., 2018).

Electronic cigarettes are also promoted on social media by celebrities and influencers who are sponsored by the vaping industry, contributing to users’ positive attitudes towards the brand, and increasing brand recognition and product loyalty (Phua and Lim, 2022). Also, the vape industry uses sponsored users as an approach to reach a varied range of potential consumers (Collins et al., 2018). A related type of electronic cigarettes promotion on social media is flavour appeal, which is used to attract especially vulnerable users in lower age groups (Krishen et al., 2021), suggesting the urgent need for stricter marketing regulations related to e-liquids promotion, as it can cause serious risk for its consumer’s health (Marques, Piqueras and Sanz, 2021).

The unregulated marketing of electronic cigarettes, including on social networking sites, appeals the most to vulnerable youths (Padon, Maloney and Cappella, 2017). Similarly observed in traditional media, the tobacco industry’s main approach to reaching adolescent audiences is associating their product with youths’ psychological and social needs (Wakefield et al., 2003). Broadly evidenced in the literature, electronic cigarette use among adolescents is strongly associated with the escalation of tobacco cigarette smoking, reflecting the significant health-related harm of vaping (O’Brien et al., 2021). Consequently, there is an urgent need to avoid resurgent tobacco use initiation due to 21^st^-century marketing practices (Padon, Maloney and Cappella, 2017).

The 13^th^ article of the WHO Framework Convention for Tobacco Control fomented the restriction of conventional cigarette promotion and advertisements (World Health Organization et al., 2009), however, electronic cigarettes, as an emerging tendency, were not included in this article (Booth et al., 2019). Despite Facebook and Instagram announcing that vaping promotion on their platforms would be banned, this self-regulation lacks information (Klein et al., 2020), suggesting the urgent need to improve marketing regulations of electronic cigarettes on social media platforms, aiming to protect especially naïve individuals (Camenga et al., 2018).

Social influence theory can be used to explain how social media can shape individuals’ perceptions and consequently behaviour towards vaping (Alpert, Chen and Adams, 20209). Social influence is defined as a change of feelings, thoughts, and attitudes from interactions with others (Walker, 2015). In a large social networking site, perceived social pressure can result in users’ confirmation of other online behaviours exposure (Lee and Hong, 2016). Social norms refer to a group of rules reflecting what individuals should and shouldn’t do based on their social environment, driving individuals’ behaviours to more acceptable social choices (Hechter, et al., 2001). Thus, electronic cigarette advertisements may promote vape products by altering people’s perceived social norms and risk perceptions about vaping (Zheng and Lin, 2021).

Furthermore, the vape industry benefits from social influence by using celebrities and social media influencers to promote and endorse their products, recognizing that through their credibility (Phua and Lim, 2022), consumers can transfer their expectations and feelings toward the advertised brand (Till and Shimp, 1998). Watching pleasant or entertaining advertising including a celebrity can activate an individual’s appetitive system and thus, facilitate electronic cigarette use intentions and vaping urges (Sanders-Jackson et al., 2019).

Electronic cigarette online exposure and social media use has been associated with increased users’ willingness to experiment with vape products and positive perceptions of vaping among diverse types of individuals smoking status, as observed in the USA and the UK (Booth et al., 2019; Sawdey et al., 2017; Pokhrel et al., 2018; Vogel et al., 2020). Despite social media being considered a large focus group (Ayers, Althouse and Dredze, 2014), allowing users to organically discuss and indirectly promote electronic cigarettes online (Allem, et al., 2017), concrete evidence should also be directed to the commercial promotion of electronic cigarettes derived from the vape industry.

As high-income countries, core anglophone nations also share a high prevalence of electronic cigarette use (Jerzyński et al., 2021), especially among youths (Hammond et al., 2019; Venkata, Palagiri and Vaithilingam, 2020; Lyzwinski et al., 2022). Core anglophone countries consist of a group of nations sharing English as the main language in addition to historical, cultural, and political attachments. Its main composition includes the UK, Ireland, the United States of America (USA), Canada, Australia, and New Zealand (Nossal, 2012).

In the USA, the first generation of electronic cigarettes was introduced in 2007 (Hiemstra and Bals, 2016), and since then, is the most used non-combustible tobacco product among youths and young adults (Murthy, 2016). An estimated 1.7 million adults or 3.7% of the American population used electronic cigarettes in 2020, reflecting the severity of the public health issue (Cornelius et al., 2022). The rise of vaping in the USA and the EVALI outbreak in 2019 were considered an epidemic (Besaratinia and Tommasi, 2020; Jones and Salzman, 2020; Farzal et al., 2019), raising public health concerns and stimulating further research and policymaking (Centers for Disease Control and Prevention, 2022).

According to a report commissioned by Public Health England in 2018, electronic cigarette experimentation among never smokers in the UK was associated with subsequent regular smoking, and despite a causal link not being established between both practices, the common liability’ hypothesis is the most reasonable justification (McNeill, et al., 2018). Despite regulations being introduced in the UK aimed to control electronic cigarette marketing across diverse channels including social media, further adjustments are needed, marketing violations and compliance issues are evidenced and the control system to regulate digital marketing is defective (Stead, et al., 2021).

Despite efforts from the US Federal Food and Drug Administration (FDA) toward a decrease in electronic cigarette promotion targeting youths, and social media restrictions attempts, online appealing content remains under regulation (Vassey et al., 2022) representing the urgent need to act towards stricter marketing policies as a key approach to reduce ENDS propagation globally (World Health Organization, 2022). In Canada, comprehensive marketing restrictions were associated with lower levels of exposure to electronic cigarettes and a lower prevalence of vaping, suggesting that regulatory policies should be fomented aiming to protect those in vulnerability (Hammond et al., 2020).

### The rationale of the review

Despite being considered a healthier alternative to conventional cigarettes and a potential aid to smoking cessation cigarettes (Hartmann-Boyce et al., 2021), the emerging popularity of ENDS especially among youths (Burt and Li, 2020) represents a major global public health concern. Therefore, the premature initiation of electronic cigarette (Bourke, Sharif and Narayan, 2021) in association with the escalation of cigarette use (Leventhal et al., 2015), reflect the paradox of vaping (Al-Hamdani and Manly, 2021). The popularity of social media among youths and the lack of marketing regulations in comparison with traditional media suggest that a better comprehension of their role in electronic cigarette use is urgently needed (Camenga et al., 2018).

The validated association of the promotion of cigarettes in traditional media on smoking behaviours especially among youths (Evans et al., 1995), contributed to long-term restrictive marketing policies (World Health Organization et al., 2009), resulting in a decrease in smokers’ awareness of pro-smoking cues (Kasza et al., 2011) and a reduction of conventional cigarettes use (Henriksen, 2012). Therefore, understanding the strategies and patterns used by electronic cigarette brands on social media can contribute to improved marketing regulations and policymakers’ decisions, aimed to decline the prevalence of vaping worldwide.

Considering the complex nature of the proposed topic, an accurate review of the literature is necessary to map the current evidence and identify the main gaps, contributing to further research on the topic.

### Aims and objective

The proposed scoping review intends to evaluate the association between electronic cigarette commercial promotion on social media and its influence on vaping initiation and positive perception among adolescents and young adults in anglophone countries, aiming to:

1. Explore the existing evidence related to the influence of electronic cigarette commercial promotion on social media on vaping initiation, positive perceptions of vaping, and subsequent use of cigarettes in anglophone countries, through a diverse range of study designs.
2. Map the existing literature to determine whether systematic reviews related to the research topic are available or feasible to be developed, or whether primary research is first required.
3. Describe, summarize, and disseminate in detail the key concepts and findings related to the research topic of interest, contributing to consumers, policymakers, and practitioners’ data accessibility.
4. Establish what prospective research must cover, by identifying the main gaps in the literature related to the research topic, contributing to a long-term, improved understanding of the correlation between commercial promotion of electronic cigarettes in social media and its impact on vaping initiation, positive perceptions of vaping and subsequent cigarette use.

## METHOD

A scoping review was chosen to address the topic based on the rationale and objective justifications of the study. According to Arksey and O’Malley (2005), conducting a scoping review is usually driven to examine the research activity of a specific topic, and to establish the value of conducting a full systematic review (Arksey and O’Malley, 2005). Scoping reviews can also be undertaken with the purpose to summarize and describe in detail the available literature and disseminate the research findings providing relevant information to policymakers and practitioners. Furthermore, a scoping review aims to identify gaps in the existing evidence base, with the potential to guide and contribute to prospective research efforts related to the topic (Arksey and O’Malley, 2005).

Differing from systematic reviews, scoping reviews are particularly valuable to summarize the existing literature from areas of emerging evidence (Peters et al., 2015) which are not comprehensively reviewed, and present a complex and heterogeneous nature (Mays, Roberts and Popay, 2001). In comparison with systematic reviews, scoping reviews tend to cover a broader topic, including diverse types of applicable study designs (Arksey and O’Malley, 2005). Furthermore, scoping reviews can be conducted according to time and location relevance, such as specific countries or contexts (Anderson et al., 2008). Considering the commercial promotion of electronic cigarettes on social media platforms in core anglophone countries and the nature of scoping reviews, the adopted methodology is suitable for the proposed topic, as it englobes a broad and diverse new study areas, including vaping behaviour, multiple social media platforms, international coverage, and marketing regulations. With no identified systematic review addressing the research question, secondary research is needed to identify the existing literature gaps, and where there is substantial and sufficient evidence to conduct a full systematic review.

A scoping review was selected as the main method for conducting the research, as they can provide in a condensed time (in comparison with full systematic reviews), a transparent method to map relevant evidence (Arksey and O’Malley, 2005). To conduct the scoping review, the study will follow Arksey and O’Malley’s framework underpinned by five methodological steps (outlined in Table 1 below), developed iteratively and reflexively, ensuring the literature was covered accurately (Arksey and O’Malley, 2005). In addition to the Arksey and Malley framework for scoping reviews, this study will also be guided by recommendations provided by Levac and collaborators, to enhance a team-based approach to scoping reviews methodologies (Levac, Colquhoun and O’Brien, 2010).

**Table 1.** Arksey and O’Malley’s (2005) framework for conducting a scoping review (Arksey and O’Malley, 2005).

| STAGE | DESCRIPTION |
| --- | --- |
| *Stage 1* | Identifying a research question |
| *Stage 2* | Identifying Relevant Literature |
| *Stage 3* | Select Studies |
| *Stage 4* | Charting the data |
| *Stage 5* | Summarize, synthesize, and report the results |

The scoping review will be also reported according to the Preferred Reporting Items for Systematic Reviews and Meta-Analyses - Scoping Review (PRISMA-ScR) (Tricco et al., 2018).

### Identifying a research question

Establishing a clear and well-structured framed question is a crucial stage in conducting a scoping review (Arksey and O’Malley, 2005), and depends on the investigator’s ability to identify, based on their practices and experiences, a leading research question, which will define the relevance and suitability of the topic of interest (Aslam and Emmanuel, 2010). Broadly used in systematic reviews, PICO (Population, Intervention, Comparison, Outcome) is considered a gold standard framework for search strategies (Cooke, Smith and Booth, 2012; Cochrane, 2015).

In the proposed scoping review, the intervention element of the PICO framework is inappropriate to address the topic of interest, as it relates to clinical practices, procedures, and treatments (Aslam and Emmanuel, 2010). Therefore, an adapted framework, PECO (Population, Exposure, Comparator and Outcomes) were identified as an alternative research question formulation tool for this study (Morgan et al., 2018), as the exposure element is more coherent with electronic cigarette commercial promotion in social media. The PECO framework is useful for environmental, public, and occupational health research, and contributes to improved information sources for decision-makers (Morgan et al., 2018). The following elements were identified through the PECO framework, aiming to formulate a well-defined research question (Table 2):

**Table 2.**
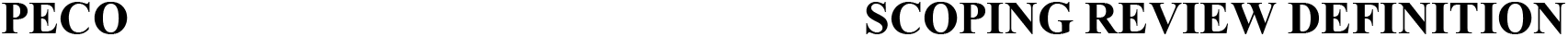
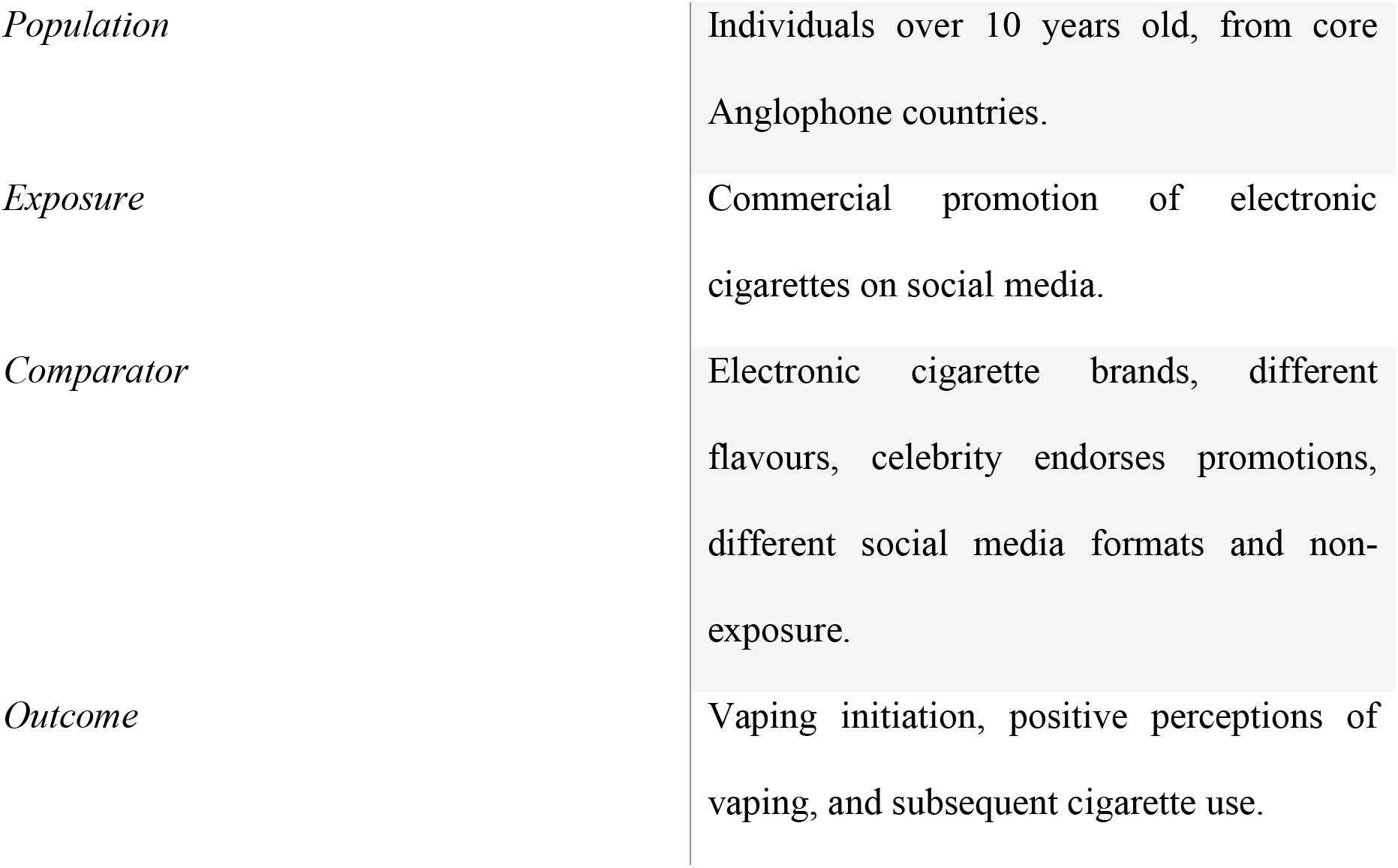
PECO framework to structure the research question.

**SCOPING REVIEW DEFINITION**
|  |  |
| --- | --- |
| *Population* | Individuals over 10 years old, from core Anglophone countries. |
| *Exposure* | Commercial promotion of electronic cigarettes on social media. |
| *Comparator* | Electronic cigarette brands, different flavours, celebrity endorses promotions, different social media formats and non-exposure. |
| *Outcome* | Vaping initiation, positive perceptions of vaping, and subsequent cigarette use. |

Based on the PECO framework guidance, the aim of the scoping review was refined into a clear and concise main research question:

1) Does the commercial promotion of electronic cigarettes on social media influence vaping initiation, positive perceptions about vaping, and subsequent cigarette use among individuals from anglophone countries?

In addition to the main research question, the following question has been also identified:

2) What further research is needed to evidence the association between the commercial promotion of electronic cigarettes on social media and vaping initiation, positive perceptions about vaping, and subsequent cigarette use among individuals from anglophone countries?

### Identifying Relevant Literature

Seven databases will be searched in this scoping review in the field of health sciences, public health, social sciences, and psychology. The databases selected underpinned their relevance to the research question and were identified in previous systematic and scoping reviews related to electronic cigarette promotion on social media (Kwon and Park, 2020; Lee et al., 2020; Amin, Dunn and Laranjo, 2020). The selected databases will be the Cochrane Database of Systematic Reviews (CDSR), Cochrane Central Register of Controlled Trials (CENTRAL), Epistemonikos, Medline, Embase, PsycInfo, and Science Citation Index (SCI). By following Cochrane guidelines (Higgins and Green, 2011), an additional search will be conducted within previous reviews on equivalent topics and reference lists will be checked to increase the study’s credibility and identify potential missing studies.

Diverse combinations of terms and their respective synonymous, variations, and abbreviations will be used in the search to identify relevant studies. Electronic cigarette terms will include electronic nicotine delivery system (ENDS), electronic nicotine, e-cigarettes, vapers, and e-vapers. Commercial promotion terms will comprise marketing and social marketing. advertisement, and direct-to-consumer. Regarding social media topics, terms will englobe social networking, internet, Facebook, Instagram, Snapchat, YouTube, and Twitter.

The terms were chosen based on the PECO question formulation framework, and from the previous search, strategies applied in topic-related systematic reviews (Kwon and Park, 2020; Lee et al., 2020; Amin, Dunn and Laranjo, 2020). Medical Subject Headings Terms (MeSH) is the list of standardized subject headings and were used in the scoping review search strategy, increasing the citations range on databases such as Medline (Dhammi and Kumar, 2014) and the Cochrane Library. No language restrictions will be applied as limiting search terms to the English language could exclude potential studies and their relevant information (Jüni et al., 2002). However, given the study population (individuals from core anglophone countries), few studies written in a non-English language are expected to be identified.

### Data Management

The search results originating from each selected database will be entered into the reference management software EndNote (Peters, 2017), where duplicated papers will be automatically identified and excluded. To increase the accuracy of the data management, a manual inspection will be conducted by the main reviewer to detect any further duplicate references, with their subsequent exclusion. After completing this stage, studies will be formatted into RIS files and will be uploaded to Covidence, a screening and data extraction software (Cleo et al., 2019), to proceed with the study selection stage.

### Study Selection

The eligibility of the studies for the scoping review will be based on the PECO framework for formulating a question (table 2), which will guide the inclusion and exclusion criteria establishment, through precise definitions of the population, exposure, comparator, and outcome of interest (Morgan et al., 2018).

#### Inclusion Criteria

Included studies must comprise individuals over 10 years old from core anglophone countries, consisting of the UK, Ireland, United States of America (USA), Canada, Australia, and New Zealand. The selection of the setting was based on the social, cultural, and historical similarities between the populations, and due to their common primary language (Nossal, 2012). Additionally, as high-income countries, core anglophone nations also share a high prevalence of electronic cigarette use (Jerzyński et al., 2021), especially among youths (Hammond et al., 2019; Venkata, Palagiri and Vaithilingam, 2020; Lyzwinski et al., 2022).

The exposure of interest of the studies must focus on the promotion of electronic cigarettes on public and accessible social media, including Instagram, Twitter, Snapchat, Facebook, and YouTube. The promotion of electronic cigarettes on these platforms derived from the vape industry (described in this study as commercial promotion), must contain the following content characteristics: URLs to retail websites, campaigns, discounts, coupons, publicities, prize draws, adverts, and paid partnerships. The content must be displayed via video (e.g., YouTube videos), photo (e.g., Instagram pictures), or text (e.g., tweets). The commercial promotion must be mediated by the vape industry, including vape brands, vape sellers, and paid celebrities/social media influencers.

Comparative studies will be eligible if assess differences between populations (e.g., England vs. Canada or adolescents vs. adults); social media (e.g., Facebook vs Twitter); electronic cigarettes brands (e.g., JUUL vs Geekvape), before and after the exposure; and with and without the exposure. The primary outcome of interest will be vaping initiation, and secondary outcomes will include a positive perception of vaping, vaping behaviour, and vaping escalation to cigarette use.

Positive perception (or sentiment) about vaping through the commercial promotion of electronic cigarettes on eligible social media can be represented through user content engagement, such as liking, commenting, and sharing reactions. Additionally, data extracted from cross-sectional and longitudinal studies including the Public Assessment of Tobacco and Health (PATH) can provide access to social media users’ perceptions of vaping through electronic cigarette commercial promotion.

The commercial promotion of electronic cigarettes on social media can be analysed and categorised into topics or themes (Sparker and Holloway, 2005), aiming to explore the patterns of the messages, hashtags, tweets, photos or videos of e-cigarette brands’ content. Therefore, content or thematic analysis studies will be eligible if provide a relevant evaluation related to the outcome of interest, such as users’ engagement.

Content analysis or thematic analysis studies will be included if they met all the following criteria:

1. Assess the commercial content of e-cigarettes on eligible social media platforms, via text, photo, or video.
2. Categorize and provide relevant analysis of commercial content derived from the vape industry.
3. Evaluate the relationship between electronic cigarette commercial content on social media with vaping initiation, and secondary outcomes, such as vaping positive perception. Positive perceptions included users’ content engagement, through likes, positive comments, and sharing.

#### Exclusion Criteria

According to the WHO, individuals under 10 years old are considered children (World Health Organization, 2022), and will not be eligible for this scoping review. Vaping initiation in paediatric individuals is a complex public health issue, with severe health risks, and specific regulations demand (Virgili et al., 2022). Since this population requires a focused assessment, they will be excluded from this prospective research. For the set criteria, any country outside the core anglosphere will be rejected.

As this proposed scoping review aims to evaluate the influence of e-cigarette commercial promotion on social media on vaping initiation and positive perceptions of vaping, studies assessing multiple exposures such as tobacco-related products (cigarettes, pipe, or cigarillo) will be excluded, intending to eliminate bias risks. Similarly, traditional media such as television, newspaper, and radio will not eligible, as this study focuses on the distinguishing features of social media platforms, due to their broad reach range and specific marketing strategies (Freeman, 2012). Dating social media such as Bumble and Tinder, retail websites, and online forums will also be excluded.

User-generated content is derived from regular social media users, and refers to non-sponsored posts, usually reflecting individuals’ personal opinions or experiences related to a specific topic, with no commercial affiliations. Studies assessing user-generated content related to electronic cigarettes will not be eligible, as this scoping review aims to study the vape industry influence as the main exposure.

Studies comparing e-cigarettes with other tobacco-related products such as cigarettes, pipes, and hookah will not be eligible as meet the exposure exclusion criteria. Equally, eligible social media with traditional media such as television, newspaper, and radio will not be eligible for comparison purposes on this scoping review. Comparison between the content of people vaping vs. regular posts will be excluded, as are not directly related to the vape industry. Vaping prevention content and vaping risk awareness content are not suitable as a comparator because they both meet exposure exclusion criteria. Differences between organic content vs. commercial content will not be adequate for this scoping review, as are not the central focus of the review purpose.

Studies providing divergent outcomes such as vaping cessation, vaping decrease, adverse effects of electronic cigarettes on health, vape risk awareness, and negative perceptions of vaping are not suitable, as are contradictory to the scoping review purpose.

Content analysis or thematic analysis studies will be **excluded** if meet one of the following criteria:

1. Assess social media organic content related to e-cigarettes, such as personal photos and forums where commercial topics are banned, as this scoping review’s main objective is related to e-cigarette promotion by the vape industry only.
2. Do not assess the promotional content of electronic cigarettes on social media platforms, including commercials, campaigns, paid adverts, and advertising mediated by the vape industry.
3. Only provides a narrative description of the promotional content of e-cigarettes on social media platforms, without evaluating their impact on vape initiation and positive perceptions of vaping.

### Data management

Covidence, a computer software, will be used to support the study selection process, contributing to a more streamlined workflow (Kellermeyer, Harnke and Knight, 2018). Apart from the main author, independent reviewers will be invited to work on the platform to decrease the risk of bias during the screening and extraction stages. Settings will be configured to provide highlights of exclusion and inclusion criteria established through the PECOS framework, and exclusion criteria will be numbered in priority order to cover studies with multiple exclusion justifications.

To identify all potential studies related to the research question, the title and abstracts will be initially screened. If eligible and mutually approved after an independent assessment by independent reviewers, papers will be subsequently included in the full-text review stage. Once full texts are reviewed by independent reviewers, those meeting all the inclusion criteria will be extracted for scoping review purposes. In both stages, papers with challenging classification facing decision conflicts will require a mutual consensus from the reviewers, consequently increasing the study’s accuracy. Additionally, to increase the study transparency, studies will be displayed through the PRISMA flowchart diagram.

### Extracting the data

The fourth stage of Arksey and O’Malley’s framework for scoping reviews refers to an analytical method of charting or mapping the data from included studies, also known as data extraction (Arksey and O’Malley, 2005). Although a standardised data extraction is not delineated in Arksey and O’Malley’s framework, additional guidance and recommendations were considered, pondering the specific challenges for the data extraction methodological framework stage, and proposing respective refinements (Levac, Colquhoun and O’Brien, 2010).

Aiming to create a descriptive summary for all the studies selected for inclusion, customized data charting tables will be formulated according to the research question specificities and are displayed in Table 3 and Table 4. An iterative, collaborative, and process-oriented data process will be followed during this stage (Levac, Colquhoun and O’Brien, 2010). A descriptive summary of the extracted studies template is displayed in Table 3 below, including general bibliographic information, population description, assessed social media, study design, conflicts of interest and funding of included studies.

**Table 3.** Template of the descriptive summary of extracted studies:

| **CATEGORY** | **DESCRIPTION** | **HEADING** |
| --- | --- | --- |
| *Biography information* | Studies biography information, including author(s) name and year of publication | Author and year of publication |
| *Population description* | Population characteristics and setting where the study was conducted | Population |
| *Social networking sites* | Social media assessed | Social media |
| *Study design* | Study design | Study design |
| *Conflicts of interest* | Any conflict of interest of the authors with the study | Conflicts of interest |
| *Funding* | Any provider source for conducting the studies | Funding |

**Table 4.**
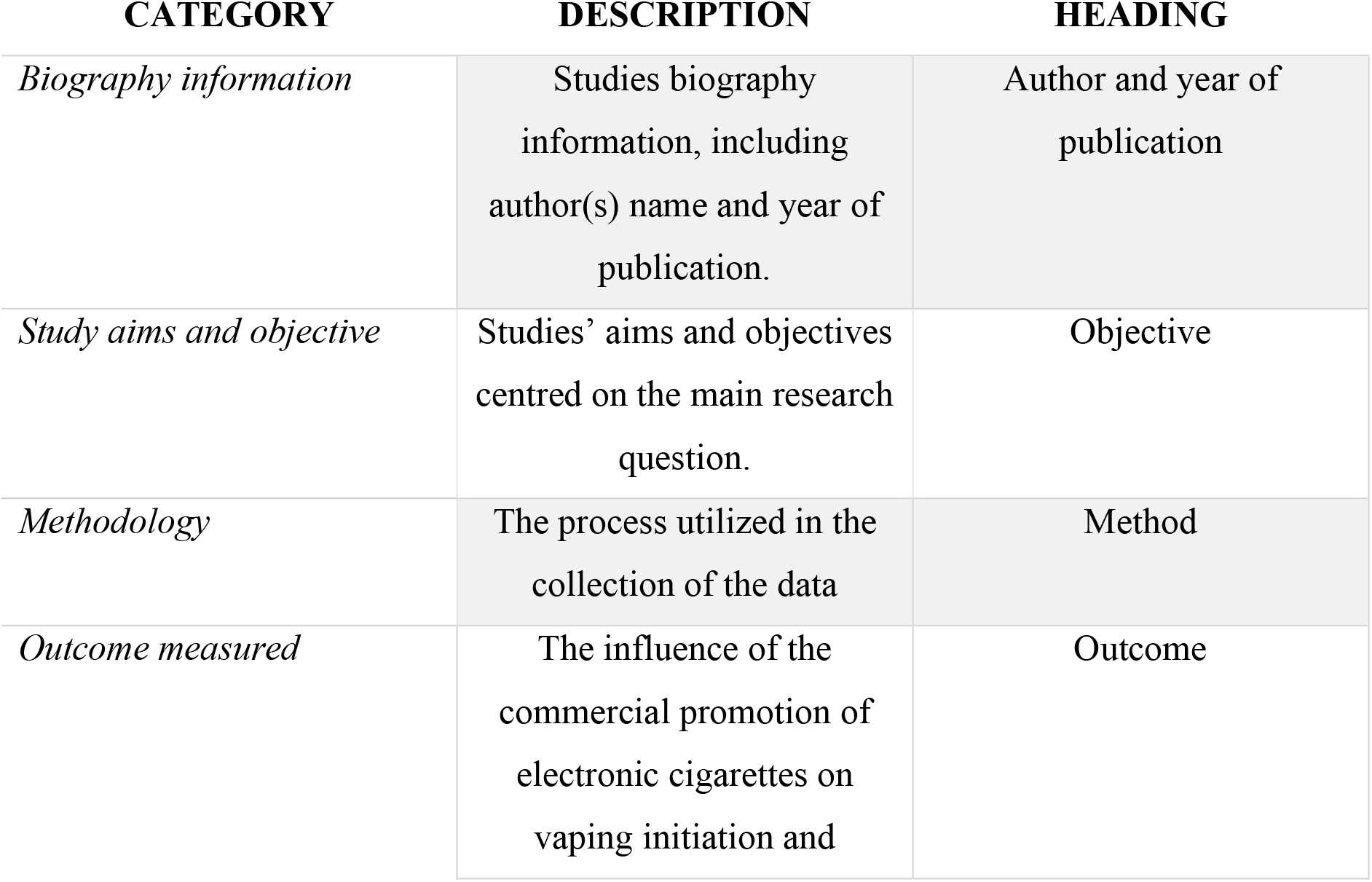
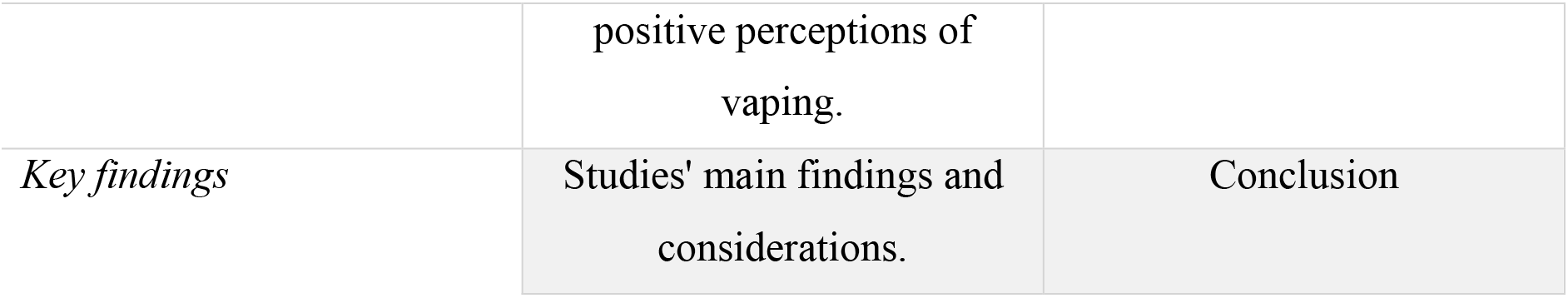
Second template of the descriptive summary of extracted studies:

In addition to the first data chart, a second descriptive summary template of the extracted studies template is displayed in Table 4 below and includes the study’s general bibliographic information, objectives, method, outcomes, and conclusion of the included studies.

### Collating, summarizing, and reporting results

According to Arksey and O’Malley’s framework, the final stage of the scoping process is based on the reviewer’s ability to summarize, synthesize, and report the results retrieved from the included studies (Arksey and O’Malley, 2005). Despite being the most extensive step of the scoping review, it lacks details. Therefore, by following a more refined framework developed by Levac and collaborators, the last phase was divided into three systematic and meaningful steps, contributing to reporting the findings with a more rigorous methodology. The three steps consist of an analysis of the data, reporting the results, and applying meaning to the results (Levac, Colquhoun and O’Brien, 2010), which are described in detail in the following sections.

#### Step 1 - Analysis of the data

As described in the original scoping review framework proposed by Arksey and O’Malley, the data analysis phase should include a descriptive numerical summary and a thematic analysis of the included studies. Therefore, the descriptive summary will be essential to display relevant information and characteristics of the included papers, comprising, for instance, their respective nature, design, population descriptions, geographic distribution, and outcomes of interest (Arksey and O’Malley, 2005).

The next element of this stage refers to the thematic analysis, which development usually resembles a qualitative data analytical technique (Levac, Colquhoun and O’Brien, 2010). With the main objective to break the data into relevant topics, the thematic analysis will contribute to the identification of patterns within the extracted data and was conducted using previously defining themes according to the research question (Braun and Clarke, 2017).

Thus, the thematic analysis is simply expressed by organizing the data according to pertinent themes (Arksey and O’Malley, 2005). Differently from systematic reviews, an assessment of the bias risk of the included studies will not be conducted and reported in the study, as is not a requirement for scoping reviews. Despite the purpose of scoping reviews being limited to mapping the evidence instead of critically judging the quality of the data (Arksey and O’Malley, 2005), considerations regarding the strengths, limitations, and gaps of the included studies will be reported within the results.

#### Step 2 - Reporting the results

To accurately report results, an appropriate format should be selected to inform the readers about the outcomes of each selected study, underpinned by the main purpose of the scoping review (Levac, Colquhoun and O’Brien, 2010). Considering the available guidance, this stage will be conducted through a narrative synthesis of the portrayed evidence, by assessing the descriptive summary, defined themes, and considerations of included studies. This process aims to correlate the commercial promotion of electronic cigarettes on social media with vaping initiation, positive perceptions of vaping, and secondary outcomes of interest.

#### Step 3 - Applying meaning to the results

As a final point, a consideration of the findings from a broader perspective, and their respective implications for prospective research will be developed to advance the legitimacy of the scoping review (Levac, Colquhoun and O’Brien, 2010). Considering the research question, insights into how to address future studies related to the commercial promotion of electronic cigarettes on social media will be described, contributing in the long-term, to improved vaping marketing policies. Within this review, the findings considerations, implications, and recommendations will be described.

## Data Availability

All data produced in the present work are contained in the manuscript

## Supplementary considerations

Arksey and O’Malley’s framework endorses a sixth optional stage, the consultation exercise, in which contributors (stakeholders from national and voluntary bodies, managers and practitioners of organizations, and informant career) can provide relevant insights into the research topic and complement the study with potential extra references (Arksey and O’Malley, 2005). Despite Arksey and O’Malley’s encouragement to review the conduct of the consultation exercise, a strong justification must support the decision (Levac, Colquhoun and O’Brien, 2010). Therefore, considering the rationale of the scoping review, the consultation phase is not significant in this context.

